# The use of an Artificial Neural Network (ANN) in the evaluation of the Extracorporeal Shockwave Lithotripsy (ESWL) as a treatment of choise for urinary lithiasis

**DOI:** 10.1101/2020.08.11.20172965

**Authors:** Tsitsiflis Athanasios, Kiouvrekis Yiannis, Chasiotis Georgios, Perifanos Georgios, Gravas Stavros, Stefanidis Ioannis, Tzortzis Vasilios, Karatzas Anastasios

## Abstract

**Purpose:** Artificial Neural Networks (ANNs) are simplified computational models simulating the central nervous system. They are widely applied in medicine, since they substantially increase the sensitivity and specificity of the diagnosis, classification and the prognosis of a medical condition. In this study we constructed an artificial neural network to evaluate several parameters of extracorporeal shockwave lithotripsy (ESWL), such as the outcome and safety of the procedure.

**Materials and methods:** Patients with urinary lithiasis suitable for ESWL treatment were enrolled. An artificial neural network (ANN) was designed and a unique algorithm was executed with the use of the well-known numerical computing environment, MATLAB. Medical data were collected from all patients and 12 nodes were used as inputs (sex, age, B.M.I. (Body Mass Index), stone location, stone size, comorbidity, previous ESWL sessions, analgesia, number of shockwaves, shockwave intensity, presence of a ureteral stent and hydronephrosis). Conventional statistical analysis was also performed.

**Results:** 716 patients were finally included in our study. Univariate analysis revealed that diabetes and hydronephrosis were positively correlated to the ESWL complications. Regarding efficacy, univariate analysis revealed that stone location, stone size, the number and density of shockwaves delivered and the presence of a stent in the ureter were independent factors of the ESWL outcome. This was further confirmed when adjusted for sex and age in a multivariate analysis.The performance of the ANN (predictive/real values) at the end of the training state reached 98,72%. The four basic ratios (sensitivity, specificity, PPV, NPV) were calculated for both training and evaluation data sets. The performance of the ANN at the end of the evaluation state was 81,43%.

**Conclusions:** Our ANN achieved high score in predicting the outcome and the side effects of the extracorporeal shockwave lithotripsy treatment for urinary stones. In fact, the accuracy of the network may further improve by using larger sets of data, different architecture in designing the model or using different set of input variables, making ANNs thus, a quite promising instrument for effective, precise and swift medical diagnosis.

## Introduction

Numerous multivariate computational programs have been widely used over the past decades in medicine, mainly in the oncology field. The aim is clear, to help diagnose and stage cancers and other medical conditions in various ways, as well as estimate the prognosis of critical diseases. These clinical decision tools are utilised to categorize patients by developing patterns and envisaging outcomes given a set of ‘inputs’. These ‘inputs’ may consist of specific patients’ or disease characteristics, making the generated outcome thus more accurate than other analogous statistical procedures. These multivariate programs substantially increase the sensitivity and specificity of the diagnosis, classification and the prognosis of a medical condition, eventually (1).

Artificial Neural Networks (ANNs) are simplified models mimicking the central nervous system. They are networks with highly interconnected neural computing and the capability to react to input signals, learning to adapt to the environment. It is supported that these models offer the most promising integrated approach to constructing truly intelligent computer systems. It has been demonstrated that ANNs can be effectively used as computational processors in a variety of tasks such as speech and visual image recognition, classification, data compression, forecasting, simulation (modeling) and adaptive control. They demonstrate desirable characteristics absent in conventional computing systems, such as high performance when associated with noise or incomplete input standards, high error tolerance, high parallel computing rates, generalization and adaptive learning. A typical network consists of one set sensory units that form the input layer, one or more hidden layers with computational connections and an output layer with calculation nodes (2).

Urinary lithiasis is a common problem worldwide with an increasing trend due to climate change, lifestyle modifications and diet (3). Most patients with stone disease have identifiable risk factors, it is noticeable though that there is a high stone recurrence rate, reaching approximately 50% at 10 years and 75% at 20 years (4). Therapy may be quite problematic sometimes, needing for reapeting maneuveurs, either non-invasing or invasing, with increasing risk of medical complications. Among the therapeutic alternatives, extracorporeal shockwave lithotripsy (ESWL) is a non-invasive method and represents the first choise for urinary lithiasis under specific conditions. The outcome of the ESWL depends on several parameters, such as stone location, stone size, stone composition, BMI, etc., making quite this therapeutic option quite challenging (5).

So far, different statistical models have been used to evaluate ESWL as a therapy procedure with inconclusive results. Since ESWL is a non invasive technique, the propability of discintegrating a stone avoiding side effects is the cornerstone of all mathematical or computational methods. In this study we constructed an artificial neural network to evaluate several parameters of ESWL, such as the outcome and safety.

## Materials and Methods

The database of the ESWL Department of the Univesrity Hospital of Larissa, Greece, was used for constructing the ANN. 716 concecutive, patients, 404 males and 312 females, with renal and ureteral stones that were treated by ESWL entered out study. All patients met the lithotripsy criteria, ie, a single stone less or equal than 1.5 cm in the maximal diameter, no anatomical abnormalities and no signs of urinary track infection.

ESWL was performed using the electromagnetic Dornier lithotripter SII (EMSE 220 F-XP) under fluoroscopic or ultrasonographic guidance, as previously described (6). All parameters of each session were recorded for each case, following the standard lithotripsy protocol. Ultrasonography or CT urography (CTU) prior to SWL were used to exclude patients with anatomical abnormalities. Analgesia was applied when needed (fentanyl citrate, 0.05 mg, iv).

### Statistical Analysis

Tables 1 and 2 show patients characteristics and statistical analysis. All variables were assumed to be discrete, categorical due to the group of data that was limited per category (not infinite). The chi-square test was used to check categorical variables, with p<0.05 considered as statistical significant (SPSS 25.0).

**Table 1:** Characteristics of 716 patients treated with ESWL

| Patient category | Complications<br>after ESWL | No complications<br>after ESWL | p-value |
| --- | --- | --- | --- |
| Sex:<br>Male<br>Female | 213<br>156 | 191<br>156 | 0,470 |
| Age:<br>30 or younger<br>31- 45<br>46 - 60<br>older than 61 | 18<br>76<br>136<br>139 | 29<br>69<br>117<br>132 | 0,945 |
| B.M.I.:<br>less than 18,5<br>18,5 - 24,99<br>25 - 29,99<br>30 or more | 0<br>104<br>167<br>98 | 3<br>92<br>166<br>86 | 0,035 |
| Stone location:<br>right kidney<br>left kidney<br>bladder<br>left ureteral<br>right ureteral | 131<br>116<br>7<br>49<br>66 | 113<br>121<br>8<br>58<br>47 | 0,311 |
| Stone size (diameter):<br>6mm or less<br>7 - 9<br>10 - 11<br>12 -13<br>14 -15<br>16 - 20<br>21- 32 | 42<br>89<br>78<br>59<br>44<br>47<br>10 | 31<br>78<br>71<br>67<br>40<br>51<br>9 | 0,541 |
| Comorbidity:<br>no symptoms<br>1 symptom<br>2 or more symptoms | 245<br>68<br>56 | 215<br>71<br>61 | 0,533 |
| Previous ESWL<br>sessions:<br>Yes<br>No | 262<br>107 | 146<br>201 | <0,001 |
| Analgesia:<br>Yes<br>No | 16<br>353 | 31<br>316 | 0,013 |
| Number of shocks:<br>2500 - 3500<br>more than 3500 | 349<br>22 | 327<br>20 | 0,118 |
| Intensity: |  |  |  |
| less than 40% | 3 | 2 | 0,060 |
| 40% - 60% | 78 | 52 |  |
| 61% - 80% | 265 | 267 |  |
| 81% - 100% | 23 | 26 |  |
| Pig-Tail existance: |  |  |  |
| Yes | 80 | 78 | 0,797 |
| No | 289 | 269 |  |
| Hydronephrosis: |  |  |  |
| Yes | 74 | 137 | <0,001 |
| No | 295 | 210 |  |

**Table 2:** Statistical analysis of input variables

| Variables | Mean | SD |
| --- | --- | --- |
| Age | 54,70 | 14,19 |
| B.M.I. | 27,66 | 4,25 |
| Stone location | 2,45 | 1,48 |
| Stone size (diameter) | 11,50 | 4,45 |
| Number of shocks | 3050,54 | 484,92 |

### ANN philosophy

A neural network is a sequence of ‘neurons’ organized in connecting layers (7). The structure of neural networks are formed by an ‘input’ layer, one or more ‘hidden’ layers, and the ‘output’ layer. The input signal is propagated through the network in the forward direction through layers. These neural networks are commonly referred to as multiple layer perceptrons (MLPs) and have been successfully implemented to solve difficult and varied problems through their education in a supervised way, using algorithms, known as error back-propagation algorithms. These algorithms are based on the error-correction learning rule and can be regarded as a generalization of an equally widespread adaptive algorithm filtering, called least-mean-square (LMS) algorithm. The process of error back-propagation consists of a forward and a backward passage through the different layers of the network. In the forward passage, an activity pattern (input path) is applied to the sensing connections of the network and its action is transmitted through the layers; eventually an output is produced.

During the forward passage synaptic weights are all fixed. On the other hand, during the backward passage, the synaptic weights are all adjusted according to the error-correction rule. In particular, the real network response is subtracted from the anticipated response (target) to deliver the error signal. Then, this error signal propagates back through the layers to the opposite path of the synaptic nodes (error back propagation). The synaptic weights are adapted in such a way to bring the actual network response closer to the desired response (8, 9).

### ANN structure

A high-level programming language and an environment for numerical computation and visualization was developed for a feed forward error back-propagation neural network using MATLAB. Data from 716 patients were divided into two sets: a training set of 549 patients and an evaluation set of 167 patients, intending to maintain equal frequency of outcomes in each set. A group of 12 variables, according to patients’ characteristics, defined as input variables (Figure 1).

**Fig 1:**
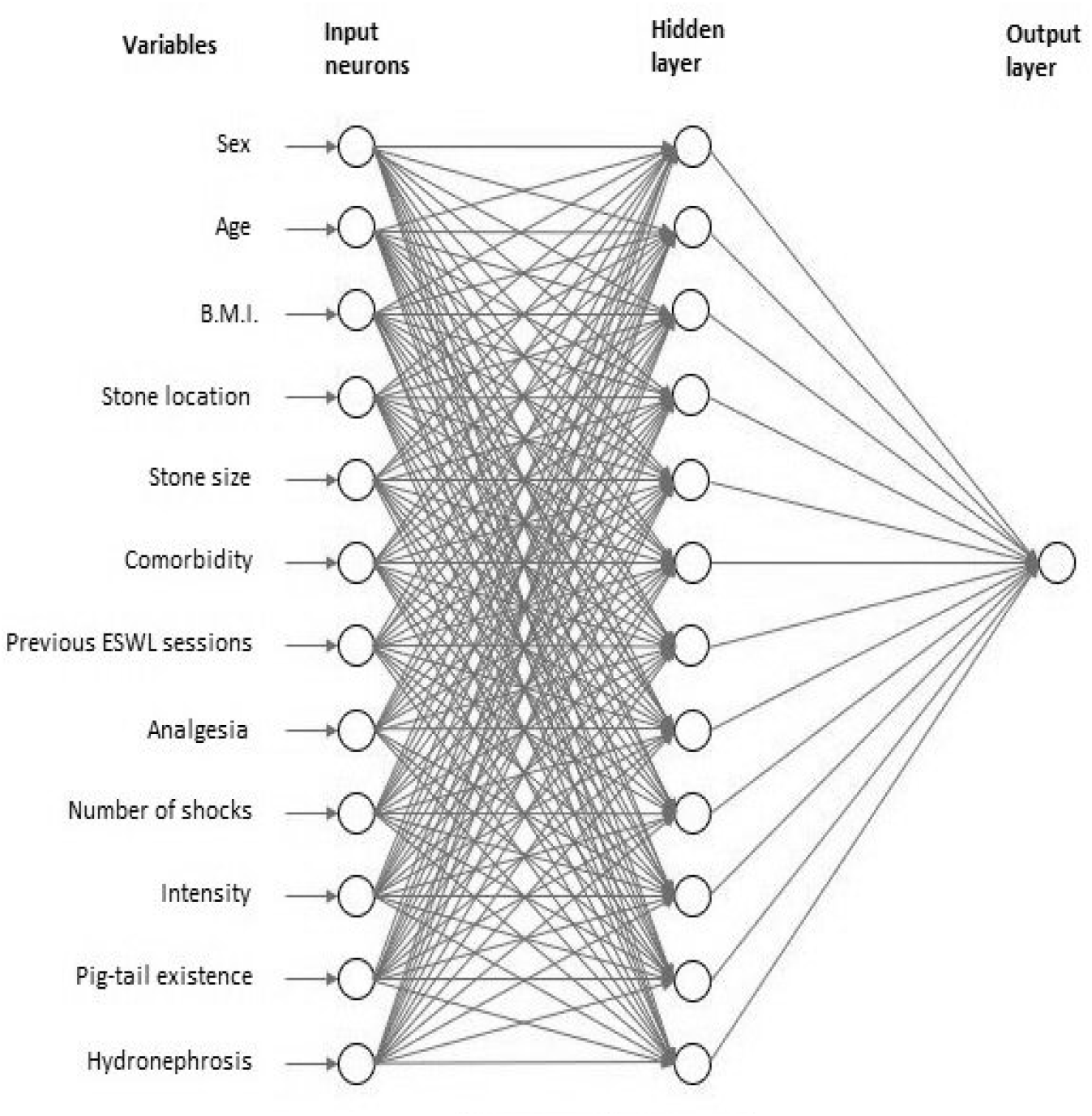
ANN nodes & connections.

Variables such as age, B.M.I or stone size contribute to the input layer with their initial values. Other variables as intensity, stone location or analgesia had the value of 1 when the category was present and 0 otherwise. “Sex” variable had the value of 1 for female patients and 0 for male. Due to the values of each variable, the input layer of ANN had 20 neurons in maximum (Table 3). Figure 2 shows how stone location has been labeled with binary values and, as a result, the variable split into 5 neurons in input layer. Each neuron corresponds to one organ in the renal system. For example, if a stone exists in right kidney, we set the value at (1,0,0,0,0). The same method was followed for comorbidity.

**Table 3:**
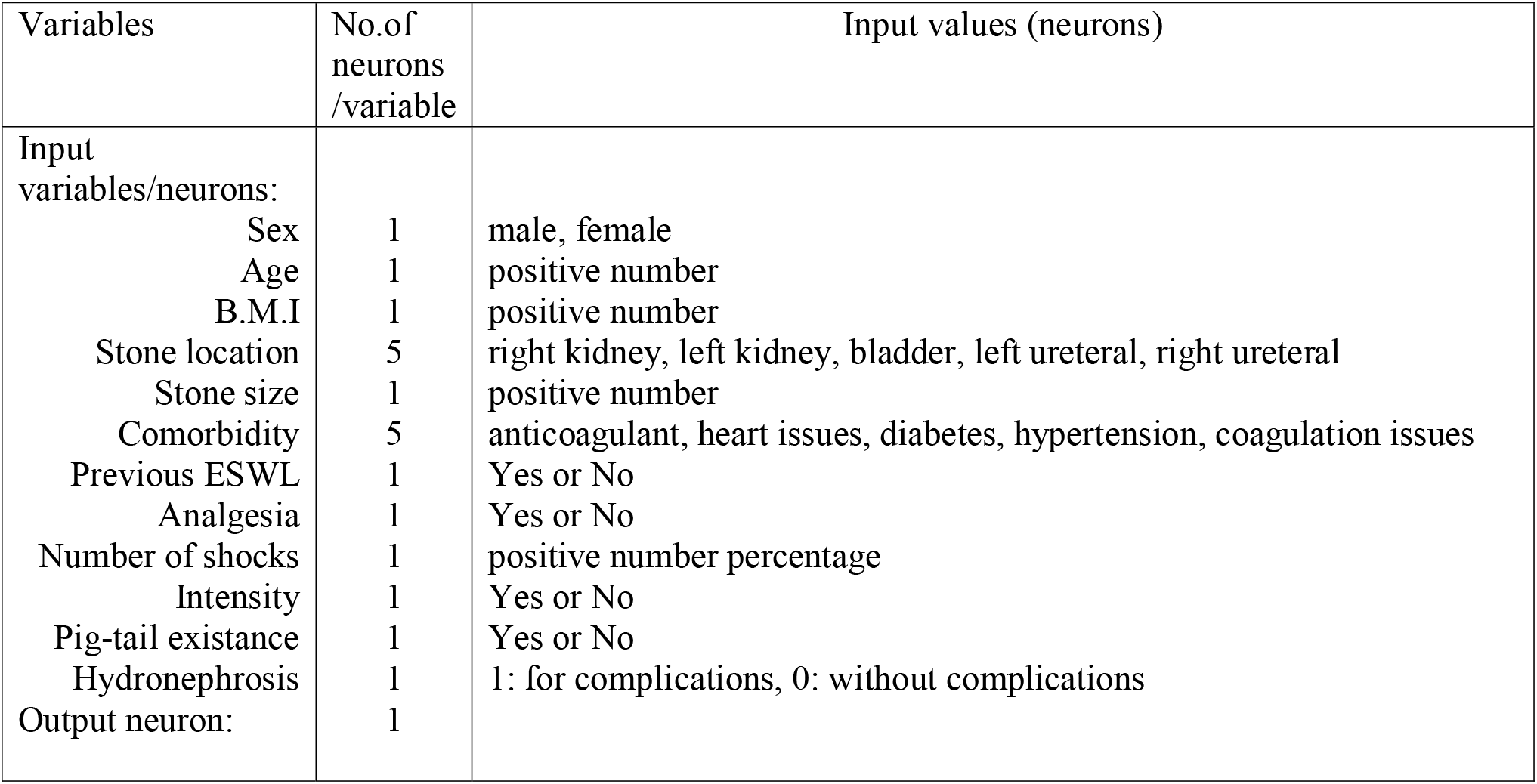
ANN input data

| Variables | No.of<br>neurons<br>/variable | Input values (neurons) |
| --- | --- | --- |
| Input<br>variables/neurons: |  |  |
| Sex | 1 | male, female |
| Age | 1 | positive number |
| B.M.I | 1 | positive number |
| Stone location | 5 | right kidney, left kidney, bladder, left ureteral, right ureteral |
| Stone size | 1 | positive number |
| Comorbidity | 5 | anticoagulant, heart issues, diabetes, hypertension, coagulation issues |
| Previous ESWL | 1 | Yes or No |
| Analgesia | 1 | Yes or No |
| Number of shocks | 1 | positive number percentage |
| Intensity | 1 | Yes or No |
| Pig-tail existance | 1 | Yes or No |
| Hydronephrosis | 1 | 1: for complications, 0: without complications |
| Output neuron: | 1 |  |

**Fig 2:**
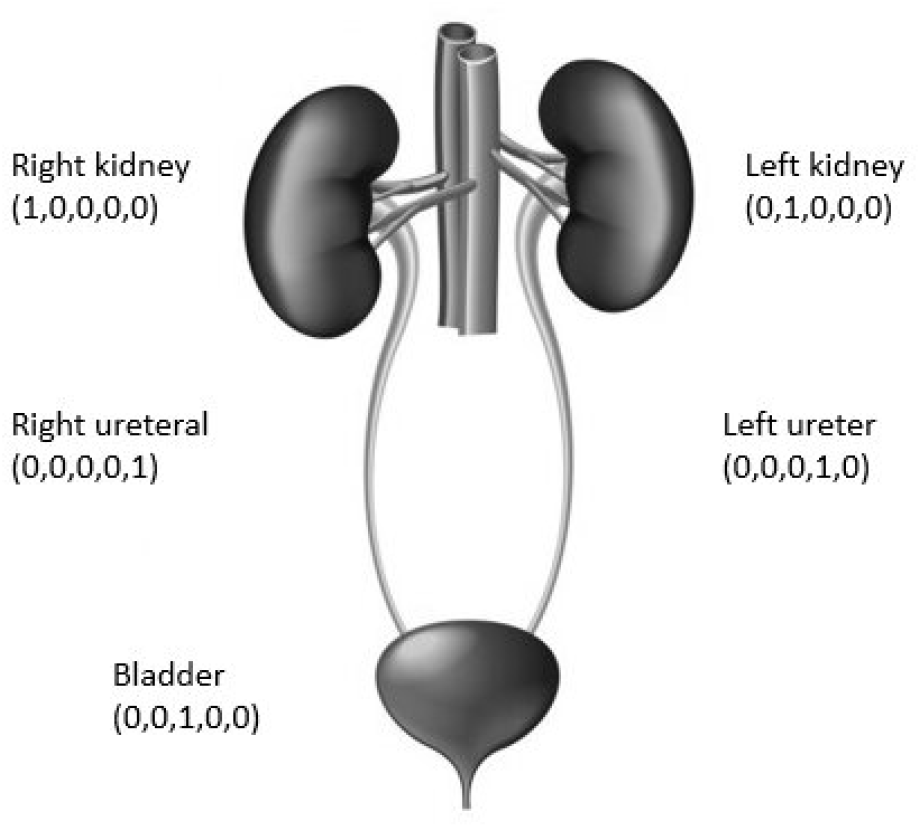
Stone location.

After the training process and careful evaluation of the network results, the hidden layer consisted of 20 neurons, giving the best network performance. The output layer consisted of 1 neuron, giving the value of 1 when complications were present and the value of 0 when there were no complications. Transfer function (from layer i to layer j) and training function were the default functions of MATLAB.

All participants were informed and gave their consent. The study was approved by the Ethics Committee of the University of Thessaly.

## Results

In all 716 patients efficacy and complications of the ESWL were evaluated in a univariate and multivariate analysis, for all the known parameters that affect the lithotripsy treatment. Univariate analysis revealed that diabetes and hydronephrosis were positively correlated to the ESWL complications, whether previous therapies and analgesia were not found to lead to any side effect. When adjusted for sex and age, multivariate analysis confirmed these results.

Regarding efficacy, univariate analysis revealed that stone location, stone size, the number and density of shockwaves delivered and the presence of a stent in the ureter were independent factors of the ESWL outcome. This was further confirmed when adjusted for sex and age in a multivariate analysis.

Initially, a data set of the statistically significant variables, as shown from the univariate and multivariate analyses, were chosen to built the neural network. 7 variables (stone location, stone size, the number and density of shockwaves delivered, the presence of a stent in the ureter, age and sex) were used as inputs in a subset of patients, giving excellent outcomes in the training of the network, but poor outcomes in the evaluation (Table 4). When all 12 parameters were used though, the outcome was improved in both the training and the evaluation of the neural network (Table 5).

**Table 4:** ANN with 7 inputs

|  | Training set<br>(334 patients) | Evaluation set<br>(84 patients) |
| --- | --- | --- |
| Performance | 92.81% | 59.52% |
| Specificity | 93.41% | 55.93% |
| Sensitivity | 92.21% | 68% |
| Positive predictive value | 92.3% | 80.48% |
| Negative predictive value | 93.33% | 39.53% |

**Table 5:** ANN with 12 inputs

|  | Training set<br>(334 patients) | Evaluation set<br>(84 patients) |
| --- | --- | --- |
| Performance | 99.1% | 75% |
| Specificity | 99.4% | 71.18% |
| Sensitivity | 98.8% | 84% |
| Positive predictive value | 98.8% | 91.3% |
| Negative predictive value | 99.39% | 55.26% |

A data set of 12 variables was finally applied to construct the ANN (fig1). The performance of the ANN (predictive/real values) at the end of the training state reached 98,72%. The four basic ratios (sensitivity, specificity, PPV, NPV) were calculated for both training and evaluation data sets (Table 6).

**Table 6:** Final ANN

|  | Training set<br>(549 patients) | Evaluation set<br>(167 patients) |
| --- | --- | --- |
| Performance | 98.72% | 81.43% |
| Specificity | 98.88% | 74.02% |
| Sensitivity | 98.56% | 87.77% |
| Positive predictive value | 98.52% | 83.82% |
| Negative predictive value | 98.92% | 79.79% |

The ANN showed high accuracy in predicting complications (81,43%) in evaluation set with high positive predictive value (83,82%), indicating that prediction of complications with the use of a neural network is very likely and extremely promising. Table 7 presents the weights that were calculated from neurons in hidden layer to output layer. Numbers show the weight of each variable in contributing to the output. These “contributions” are the weights in the ANN. A higher weight of a variable tells the ANN that it is more important than other features.

**Table 7:** Relative weight values from neurons in hidden layer to the output layer of ANN

|  |
| --- |
| -0,139862992 |
| 0,210890851 |
| 53,82936324 |
| 60,66050598 |
| 83,46458426 |
| -67,10492626 |
| -31,31098407 |
| 4,681810993 |
| -51,22454481 |
| 0,661188773 |
| -41,08985477 |
| 49,84592143 |
| -0,416188278 |
| 55,87248888 |
| -0,141036303 |
| 51,52385436 |
| 29,25816254 |
| -19,82752646 |
| -58,5080439 |
| -41,25440371 |

The input given the greatest weight by the artificial neural network was the stone location. BMI was in fourth position in terms of significance. Stone size was given negative weight by the ANN, which may be due to the total number of patients in our training set or the linearity of size measurements used as inputs that did not enable the program to locate stones with small size differences.

## Discussion

Computational intelligence methods are gaining appreciation more and more nowadays in the medical field. With a variety of paradigms, comprising expert systems, artificial neural networks, fuzzy systems, etc., these approaches are oriented in solving medical problems resistant to conventional computing methods. ANNs in particular, typically examine a relationship between the variables of a data set, which is not clearly understood. With specific variables as inputs and outputs, the network is trained choosing cases from a data set, while others are held to be used for testing the trained network. The trained network’s effectiveness then is evaluated by giving it input values from the withheld cases of the data set, which are then compared with the corresponding actual values from the testing cases. A good performance of the ANN indicates that the neural network has indiscriminated the pattern in the training cases, to identify it in cases it has never seen before (10).

Numerous studies nowadays compared the different statistical and neural computing methods, while others merged neural computing into statistical processes (11-14). In most of them, trained neural models have shown superiority as a predicting approach compared to statistical estimation tools. While the latter usually offer concise descriptive outcomes, neural networks make fewer mistakes, uncovering relationships in data sets which conventional statistics fail to identify at all. It is suggested that the backpropagation training method for artificial neural networks is equivalent to maximum likelihood estimation, making eventually the multilayered feedforward neural network a powerful modeling instrument. Furthermore, a two-layer feedforward neural network with an adequate amount of hidden nodes can estimate any continuous function accuracately. ANNs are backpropagated neural networks, which typically employ training methods (phases) to minimize errors (15).

In the latter body of evidense, ANNs for the analysis of medical data are increasing attention in the literature. Indeed, the nature of medical conditions and the complexity in differential diagnosis are critical motivators of this concern. Numerous studies have adopted this method in almost every medical field. In 1991, Baxt created a neural network for diagnosing myocardial infarction (16). Input variables used were clinical symptoms (chest pain), history and laboratory of adults addressed the Emergencies. The diagnostic accuracy was markedly improved. Karabulut and Ibrikçi applied ANNs in the diagnosis of coronary artery disease (CAD), reaching 91.2% accuracy (17), while age, cholesterol or arterial hypertension have been used as data in ANNs to diagnose CAD (18). Neural models have also been employed in other heart diseases, such as heart valve defects (19) and arrhythmias with 95% and 99.2% to 99.8% accuracy, respectively (20). In 1993, McGonigal et al. constructed a neural network model to estimate survival in patients with penetrating trauma (21), improving sensitivity over the TRISS and ASCOT survival prediction tools (22).

Another field with whidespread use of ANNs is oncology. They were initially adopted for breast and ovarian cancer in 1994, raising an argument on the suitability of certain data as inputs for ANN analysis, such as demographic, radiological, oncological and biochemical data (23). In radiology, ANNs aim at developing automated decision supporting systems, with extended application in various fields (24).

In urology, ANNs have been applied mostly in oncological diseases, such as prostate cancer (CaP). Snow et al. constructed a neural model for prostate cancer using prostate biopsy results and patient outcomes after prostatectomy. They revealed 87% and 90% accuracy in predicting biopsy results and tumour recurrence, respectively (25). Finne et al. compared a neural network based on %fPSA to conventional statistical analysis (logistic regression) or %fPSA alone, showing that the precision in predicting CaP in 656 patients who had undergone biopsy was higher in the ANN group than that of the logistic regression group (P <0.001) (26). Accordingly, Babaian et al. constructed an ANN to detect prostate cancer in 151 biopsied men. The study showed higher specificity at 92% sensitivity than the %fPSA biomarker (62% vs 11%). Interestingly, they found that, 64% of all biopsies (71 of 114 men without cancer) could have been avoided using their neural network model (27). Another ANN assessed cancer risk in regard to the outcome of prostate biopsies in 928 patients employing serum PSA, %fPSA, age, prostate volume and digital rectal examination (DRE), as inputs. At 90% sensitivity, the neural model performed better than serum PSA alone (28). Additionaly, ANN was superior when evaluated in a multicentre study with 1,188 patients (29). In final, several studies addressing neural networks exist in the literature in different medical fields, such as oncology, radiology and cardiology (30), but they can also be found in interesting conditions, ie., auditory brainstem response (31), sleep classification in infants (32), glaucoma (33), and even interhospital transport mode (34).

Our study is the first attempt in the literature to construct a neural network predicting urinary lithiasis treatment. The initial concept was to feed the network with as many data (inputs) considered statistically significant with the conventional statistical methods, as possible. It is of note that, when univariate and multivariate analyses were used, the significance of their results did not have a positive effect on the network. The outcome in the evaluation arm of the ANN was rather disappointing (59.52% performance). When all 12 parameters were used as inputs though, the performance of the network in the evaluation arm improved dramatically (75%), indicating that the algorithm used for the ANN delivers better with a greater number of inputs. Eventually, the performance of our ANN reached 81.43% in our study population.

Study limitations are the relatively small sample of patients and the lack of knowing the stone composition prior to ESWL. Yet, when a predicting tool regarding the outcome of the ESWL is constructed, only speculations can be made on the stone substances, since there is no established pretreatment method to distinguish between the different types of stones. Thus, this limitation gives strength to our ANN, since it can be used as a prediction method to all stones regardless the unknown stone composition.

In conclusion, the use of a neural network appears to be a powerful modeling tool for the diagnosis and treatment of several medical conditions. Our ANN achieved high score in predicting the outcome and the side effects of the extracorporeal shockwave lithotripsy treatment for urinary stones. In fact, the accuracy of the network may further improve by using larger sets of data, different architecture in designing the model or using different set of input variables, making ANNs thus, a quite promising instrument for effective, precise and swift medical diagnosis.

## Data Availability

The data are available by last author

